# Combinations of PCR and isothermal amplification techniques are suitable for fast and sensitive detection of SARS-CoV-2 viral RNA

**DOI:** 10.1101/2020.09.23.20200055

**Authors:** Dmitriy A. Varlamov, Konstantin A. Blagodatskikh, Evgenia V. Smirnova, Vladimir M. Kramarov, Konstantin B. Ignatov

**Affiliations:** Syntol JSC, Moscow, Russia; Pirogov Russian National Research Medical University, Moscow, Russia; Shemyakin-Ovchinnikov Institute of Bioorganic Chemistry, Russian Academy of Sciences, Moscow, Russia; Vavilov Institute of General Genetics, Russian Academy of Sciences, Moscow, Russia

**Keywords:** SARS-CoV-2, COVID-19, loop-mediated isothermal amplification (LAMP), polymerase chain displacement reaction (PCDR), PCR-LAMP, hybrid virus detection technique

## Abstract

The newly identified coronavirus, severe acute respiratory syndrome coronavirus 2 (SARS-CoV-2), causes coronavirus disease 2019 (COVID-19) and has affected over 25 million people worldwide as of 31 August 2020. To aid in the development of diagnostic kits for rapid and sensitive detection of the virus, we evaluated a combination of polymerase chain reaction (PCR) and isothermal nucleic acid amplification techniques. Here, we compared conventional PCR and loop-mediated isothermal amplification (LAMP) methods with hybrid techniques such as polymerase chain displacement reaction (PCDR) and a newly developed PCR-LAMP method. We found that the hybrid methods demonstrated higher sensitivity and assay reaction rates than those of the classic LAMP and PCR techniques and can be used to for SARS-CoV-2 detection. The proposed methods based on the modern hybrid amplification techniques markedly improve virus detection and, therefore, can be extremely useful in the development of new diagnostic kits.

## Introduction

The SARS-CoV-2 pandemic started in late December 2019, in Wuhan, China (Jiang and Shi, 2020; Wu et al., 2020; Zhou et al., 2020). By August 31, 2020, over 25 million cases of SARS-CoV-2 infection have been confirmed worldwide, with a death toll over 0.8 million. One key aspect of a safe transition from lockdown is the ability to continuously test the population for the presence of SARS-CoV-2 and hence, COVID-19 infected individuals. Nucleic acid testing is the primary method for COVID-19 diagnosis. The SARS-CoV-2 virion contains a single-stranded positive-sense RNA genome, 30,000 nucleotides in length (Zhou et al., 2020). Thus, testing involves the reverse transcription (RT) of SARS-CoV-2 RNA into complementary DNA (cDNA), followed by amplification of targeted regions of the cDNA. Currently, standard molecular techniques for the detection of the virus are quantitative RT polymerase chain reaction (RT-qPCR) (Bruce et al., 2020; Chu et al., 2020; Corman et al., 2020; Garafutdinov et al., 2020; Gray et al., 2020; Yip et al., 2020; Zhu et al., 2020) and RT loop-mediated isothermal amplification (RT-LAMP) (Ben-Assa et al., 2020; Butler et al., 2020; Garafutdinov et al., 2020; Gray et al., 2020; Lu et al., 2020; Zhang et al., 2020). RT-LAMP and RT-qPCR can be performed in either a one-step or two-step assay. In the one-step assay, RT and cDNA amplification processes are consolidated into one reaction. This assay format can provide rapid and reproducible results for high-throughput analysis. The challenge of the combined reaction process is the difficulty in optimizing the RT and amplification steps as they occur simultaneously, thereby leading to lower target amplicon generation. In the two-step assay, the reaction is carried out sequentially in separate tubes. This assay format is more sensitive than the one-step assay, but it is more time- and labor-consuming and requires optimization of additional parameters. Since the release of the SARS-CoV-2 genomic sequence, many RT-qPCR and RT-LAMP assays have been developed and used for the confirmation of many cases of SARS-CoV-2 infection during the course of the pandemic (Garafutdinov et al., 2020; Udugama et al., 2020).

RT-qPCR is the “gold standard” for molecular diagnostic assays and the method predominantly employed for diagnosing COVID-19 using respiratory tract samples (Garafutdinov et al., 2020; Udugama et al., 2020). It is a robust technique that reliably detects 30 – 50 copies of SARS-CoV-2 RNA molecules (Bruce et al., 2020; Gray et al., 2020) or less (Corman et al., 2020). However, RT-qPCR is a relatively time-consuming technique, requiring approximately 1.5–2 h.

RT-LAMP is an isothermal technique that provides a good alternative to PCR-based amplification assays. LAMP was first described in 2000 (Notomi et al., 2000). It uses a strand-displacing DNA polymerase, four or six primers, and provides a very fast and specific amplification of target DNA or cDNA at a constant temperature. One of the key advantages of using LAMP-based tests to detect infectious agents is the short duration of the assay (about 30–45 min for the DNA amplification stage). During the COVID-19 pandemic, rapid diagnostic assays such as RT-LAMP have become extremely important for high-throughput screening of patients. However, in several studies describing experiments on the detection of SARS-CoV-2 RNA molecules, RT-LAMP assays demonstrated less sensitivity than corresponding RT-qPCR tests, consistently detecting only 400 - 500 copies of RNA, with sporadic detection of as little as 120 copies (Gray et al., 2020; Zhang et al., 2020). Furthermore, RT-LAMP can generate false negative results up to 20% for patients with low viral loads testing positive by RT-qPCR (Ben-Assa et al., 2020; Butler et al., 2020).

Polymerase chain displacement reaction (PCDR) (Harris et al., 2013; Ignatov et al., 2014) and the newly developed PCR-LAMP technique (described below) are hybrid methods combining PCR-based thermocycling nucleic acid amplification with isothermal amplification. PCDR is a hybrid technique of PCR with strand displacement amplification (SDA). PCR-LAMP combines a PCR-like thermocycling mode at the initial stage of amplification with an isothermal LAMP mode during the following stage. The hybrid methods combine both the high sensitivity of PCR and the fast assay rate of isothermal amplification. In this work, we demonstrate the feasibility of applying the hybrid techniques of PCR thermocycling with SDA and LAMP isothermal amplification in the detection of SARS-CoV-2 viral RNA.

## 1 Materials and Methods

### 1.1 Enzymes and reagents

Moloney Murine Leukemia Virus (M-MLV) reverse transcriptase was supplied by ThermoFisher (Carlsbad, CA, USA). Reverase (reverse transcriptase enzyme), SD HotStart DNA polymerase (10 U/µL), SD DNA polymerase (100 U/µL), and 10× SD polymerase reaction buffer were supplied by Bioron GmbH (Römerberg, Germany). dNTPs were obtained from Bioline Limited (London, UK)). Oligonucleotide primers were synthesized by Syntol JSC (Moscow, Russia). SARS-CoV-2 viral RNA (isolate—SARS-CoV-2/human/RUS/20200417_10/2020; GenBank accession number MT890462) (25,000 copies/µL) was supplied by Syntol JSC. The real-time amplification reactions were performed using a CFX96 Touch Real-Time Detection System (Bio-Rad Laboratories, Inc., Hercules, CA, USA).

### 1.2 Two-step RT-qPCDR and RT-qPCR assays

Reverse transcription (RT) reactions were performed using M-MLV reverse transcriptase according to the manufacturer’s (ThermoFisher) instructions. Briefly, the RT reaction mixtures (20 µL each) were prepared as follows: RNA template solution (12 µL) containing 20, 200, 2000, or 20,000 copies of SARS-CoV-2 viral RNA, or 50 ng of human total RNA as a negative control, were mixed with 4µL of 5× first-strand buffer, 2 µL of 0.1 M DTT, 1 µL of 10 mM dNTP mix, 1 µL of 10 µM FP1 Primer (Supplementary Table S1) and 50 U of M-MLV reverse transcriptase. The RT mixtures were incubated for 50 min at 37 °C and then 15 min at 70 °C to inactivate the reaction.

For qPCDR and qPCR amplification of cDNA, 5 µL of the RT mixtures (containing 25% of the cDNA generated) was added to 20 µL of the respective qPCDR and qPCR master mixes that contained 1× SD polymerase reaction buffer, 0.5 U/µL SD HotStart DNA polymerase, 3 mM MgCl2, 0.35 mM dNTPs (each), and 0.4× SYBR Green I intercalating dye. The qPCR master mix contained two primers: FP2 and RP2 (0.4 µM each). The qPCDR master mix contained four primers: two inner primers FP2 and RP2 (0.4 µM each), and two outer primers FP1 and RP1 (0.2 µM each). The primers are described in Supplementary Table S1. Amplifications were carried out using a Bio-Rad CFX96 PCR machine and the following protocol: initial preheating at 92 °C for 3 min, followed by 50 cycles: 92 °C (10 s), and 68 °C (30 s).

### 1.3 One-step RT-q(PCR-LAMP) and RT-qLAMP assays

RT-q(PCR-LAMP) and RT-qLAMP assay reactions (25 µL) contained either 5, 50, 500, or 5000 copies of SARS-CoV-2 viral RNA or 50 ng of human total RNA as a negative control, 1× SD polymerase reaction buffer, 10 mM DTT, 3.5 mM MgCl_2_, 0.5 mM dNTPs (each), 0.275× SYBR Green I intercalating dye, 0.4 U/µL Reverase reverse transcriptase, 4 U/µL SD DNA polymerase, and six primers (Supplementary Table S1): outer primers (F3 and B3; 0.16 μM each), inner primers (FIP and BIP; 1.6 μM each), and loop primers (LF and LB; 1.2 μM each).

The RT step of the assays was carried out as follows: 30 s at 58 °C and then 7 min at 50 °C, followed by qLAMP or q(PCR-LAMP) reactions. qLAMP was performed at 66 °C for 40 min. q(PCR-LAMP) was carried out with an initial preheating at 92 °C for 15 s, followed by 1, 2, 4, or 6 PCR cycles: 92 °C (5 s) and 66 °C (15 s), followed by isothermal amplification at 66 °C for 35 min.

### 1.4 Data analysis

All calculations were conducted with R language usage (R Core Team, 2019). Besides standard functions, several packages were used including RDML for raw amplification data manipulation (Rödiger et al., 2017), chipPCR for Cq calculation (Rödiger et al., 2015), and ggplot2 for generation of plots (Wickham, 2016).

## 2 Results

### 2.1 RT-qPCDR vs. RT-qPCR

The sensitivity of the RT-qPCDR and RT-qPCR techniques in detecting SARS-CoV-2 RNA was evaluated using the two-step assay in which the RT and cDNA amplification steps were performed in two separate reactions. SD DNA polymerase used to amplify the cDNA is a thermostable Taq DNA polymerase mutant that has strong 5’–3’ strand displacement and 5’–3’ polymerase activities. This polymerase is suitable for PCR, PCDR, and isothermal DNA amplifications (Wang et al., 2018; Shchit et al., 2017; Ignatov et al., 2014; Smith, 2017; Alyethodi et al., 2018; Lou et al., 2018). The mixes for performing either qPCR or qPCDR were identical, except for two extra outer primers added in case of qPCDR. RT-qPCDR yielded better Cq values (−ΔCq approximately 5–7 cycles) and at least a ten-fold improvement in sensitivity than that of RT-qPCR (Figure 1 and Table 1). Under the experimental conditions, the qPCDR-based test detected as few as 5 copies of SARS-CoV-2 cDNA in less than 40 cycles. In contrast, the qPCR-based test provided consistent detection only at 50 copies of viral cDNA per reaction in 38–40 cycles.

**Table 1.** qPCR and qPCDR amplification of SARS-CoV-2 cDNA. The mean quantification cycles (Cq) ± standard deviation of three replicates for the indicated cDNA copy numbers are provided. The amplifications were carried out with SD HotStart DNA polymerase using a series of SARS-CoV-2 cDNA templates with 10-fold dilutions from 5000 to 5 copies per reaction (NTC – no template control). ΔCq is the difference between the Cq of the qPCR and qPCDR for each template dilution.

| **SARS-CoV-2**<br>**cDNA copies**<br>**per reaction** | **5000** | **500** | **50** | **5** | **NTC** |
| --- | --- | --- | --- | --- | --- |
| **Cq of qPCR** | 29.61 ± 0.04 | 33.25 ± 0.07 | 35.94 ± 0.04 | NA | NA |
| **Cq of qPCDR** | 22.72 ± 0.06 | 28.09 ± 0.05 | 33.20 ± 0.09 | 37.66 ± 1.36 | NA |
| **ΔCq** | 6.89 | 5.16 | 2.74 | — | — |

**Figure 1.**
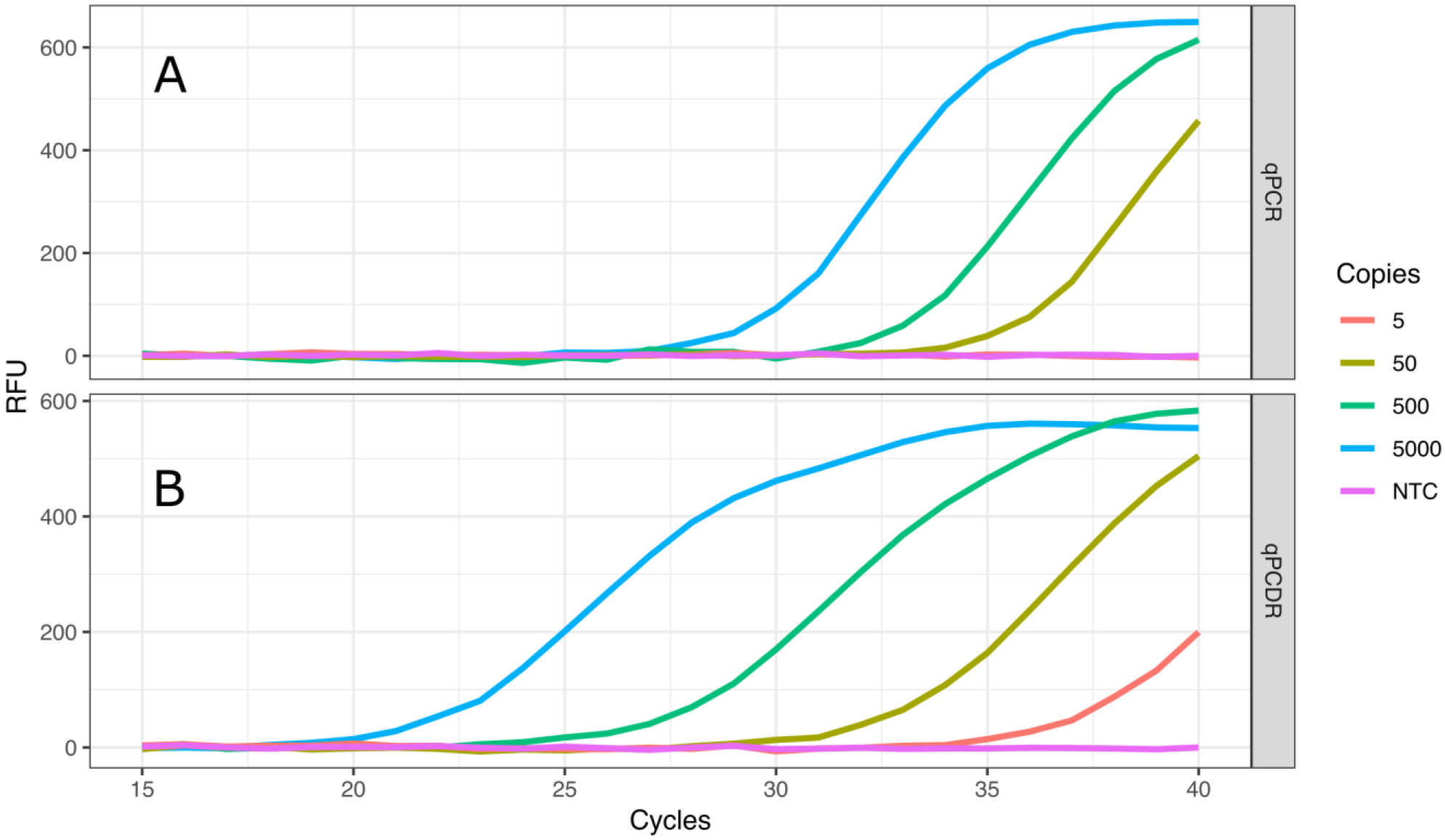
Comparison of two-step RT-qPCR and RT-qPCDR assays. After reverse transcription (RT) of SARS-CoV-2 RNA, the generated cDNA was detected by qPCR (A) or qPCDR (B). The quantitative real-time reaction assays contained 5000, 500, 50, or 5 copies of cDNA per reaction, or a no template control (NTC). RT-qPCDR provided better sensitivity in the assay than RT-qPCR. All curves reflect the mean relative fluorescence units (RFU) of three replicate reactions.

### 2.2 RT-q(PCR-LAMP) vs. RT-qLAMP

The standard RT-qLAMP and hybrid RT-q(PCR-LAMP) reactions were performed in a one-step assay, where RT and cDNA amplification were carried out in the reaction mixture containing both Reverase reverse-transcriptase and SD DNA polymerase. The standard RT-qLAMP assay was able to detect only 500 copies of SARS-CoV-2 RNA per reaction (Fig. 2 and Table 2). In contrast, the RT-q(PCR-LAMP) test provided consistent detection at levels as low as 5 copies per reaction (lower titers were not tested). Increasing the number of PCR cycles up to six before LAMP improved the assay results, but additional cycles beyond six did not provide any additional benefit (data not shown).

**Table 2.** Comparison of RT-qLAMP and RT-q(PCR-LAMP) assays. SARS-CoV-2 RNA was detected by RT-qLAMP and RT-q(PCR-LAMP) assays. The q(PCR-LAMP) amplifications were performed with 1, 2, 4, and 6 cycles of PCR. The table lists the mean quantification times (Tq) ± standard deviation of three replicates of the qLAMP and q(PCR-LAMP) reactions for the indicated RNA copy numbers. The assays were carried out with Reverase reverse-transcriptase and SD DNA polymerase using a series of SARS-CoV-2 RNA 10-fold dilutions from 5000 to 5 copies per reaction (NTC – no template control).

| **SARS-CoV-2 RNA**<br>**copies per reaction** | **5000** | **500** | **50** | **5** | **NTC** |
| --- | --- | --- | --- | --- | --- |
| **Tq (min) of**<br>**qLAMP** | 27.51 ± 0.24 | 31.47 ± 3.72 | NA | NA | NA |
| **Tq (min) of**<br>**q(PCR-LAMP);**<br>**1 cycle of PCR** | 21.30 ± 0.98 | 23.44 ± 4.27 | 28.44 ± 5.03 | 30.40 ± 3.39 | NA |
| **Tq (min) of**<br>**q(PCR-LAMP);**<br>**2 cycles of PCR** | 19.88 ± 0.85 | 21.69 ± 1.72 | 25.83 ± 5.44 | 26.86 ± 6.17 | NA |
| **Tq (min) of**<br>**q(PCR-LAMP);**<br>**4 cycles of PCR** | 18.10 ± 0.29 | 20.70 ± 0.89 | 23.31 ± 4.18 | 26.24 ± 5.06 | NA |
| **Tq (min) of**<br>**q(PCR-LAMP);**<br>**6 cycles of PCR** | 15.56 ± 0.90 | 19.54 ± 0.67 | 20.81 ± 0.76 | 22.23 ± 0.93 | NA |

**Figure 2.**
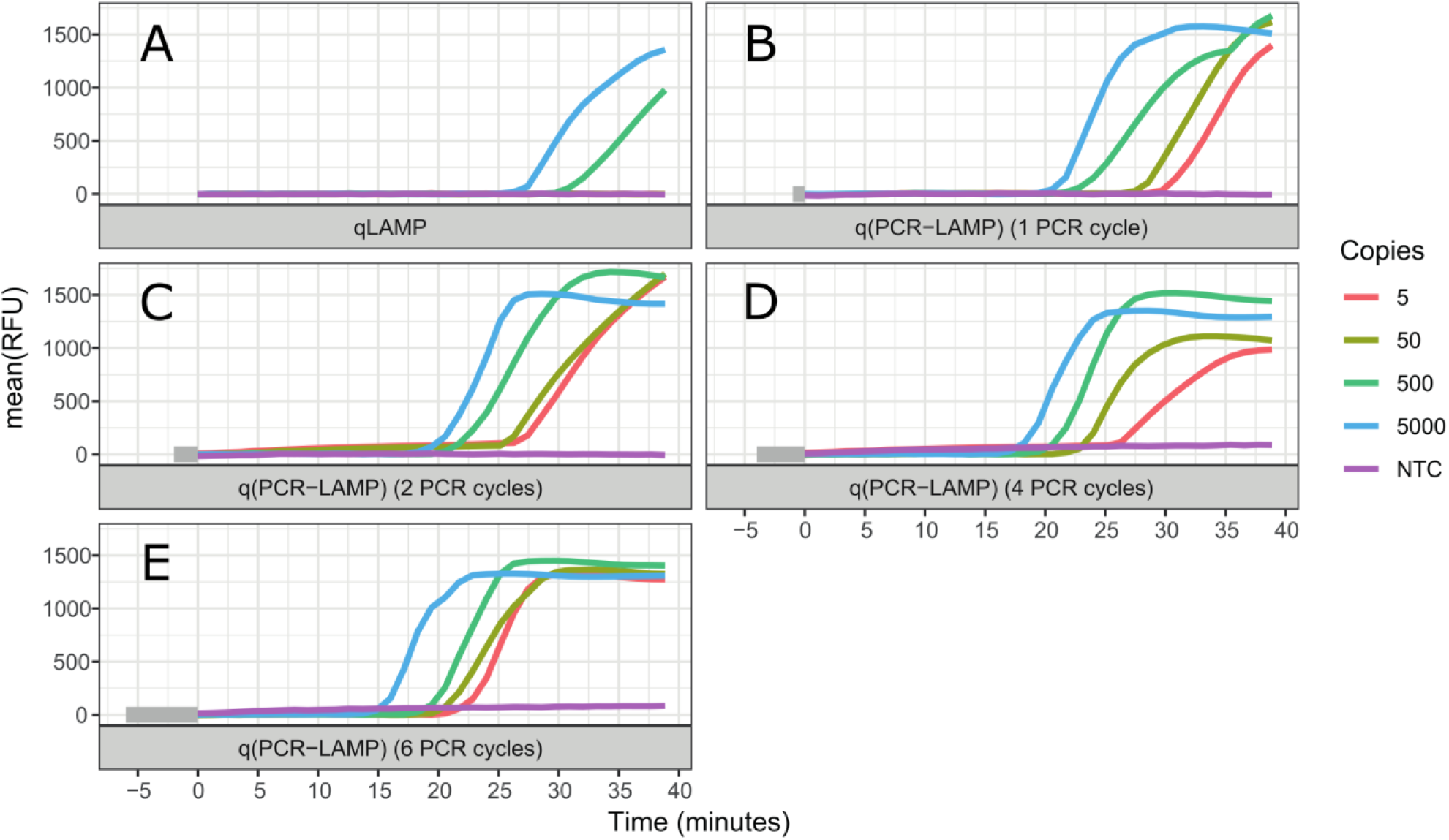
Comparison of one-step RT-qLAMP and RT-q(PCR-LAMP) assays. The reaction assays contained 5000, 500, 50, or 5 copies of SARS-CoV-2 RNA molecules, or a no template control (NTC). Negative control reactions contained 50 ng of human total RNA. The detection of viral RNA was performed by RT-qLAMP (A) and RT-q(PCR-LAMP) (B–E) assays. The RT-q(PCR-LAMP) assays were carried out with one (B), two (C), four (D), and six (E) PCR cycles before the LAMP isothermal amplification step, and provided better sensitivity than the RT-qLAMP assay. The figure shows the amplification reaction time. Time “zero” on the x-axis is the start of the isothermal (LAMP) amplification step. The time required for PCR cycling is highlighted by the gray color at the start of the timeline. All curves are the mean relative fluorescence units RFU of three replicate reactions.

## Discussion

During the COVID-19 pandemic, the availability of rapid diagnostic assays is extremely important for high-throughput screening of individuals. For the fast and sensitive detection of a target nucleic acid, we offer a new approach for DNA amplification, combining PCR and isothermal amplification techniques in one assay.

PCDR is the first method of DNA amplification, which combines PCR and isothermal amplification technique, was described in 2013. It is a hybrid of PCR and isothermal SDA. The technique requires heat denaturation of dsDNA, as in conventional PCR, as well as the strand displacement activity of DNA polymerase, as in SDA. In PCDR, at least four primers are employed and amplification is initiated simultaneously from the outer primers and the inner primers. By using a DNA polymerase with the strand displacement activity, the inner DNA strands are displaced during DNA synthesis from the outer primers and can be used as additional template strands for DNA amplification. As a result, PCDR enhances DNA amplification more than two-times per cycle as compared with the standard two-primer PCR. The mechanism and kinetics of PCDR amplification have been described previously (Ignatov et al., 2014).We used RT-qPCDR for the detection of SARS-CoV-2. The use of qPCDR allowed us to detect as few as 5 copies of SARS-CoV-2 cDNA in the reaction in less than 40 cycles, which took about 45–50 min. Under similar conditions, conventional qPCR was able to detect only 50 copies of cDNA. Thus, in agreement with earlier described findings (Wang et al., 2018; Harris et al., 2013; Ignatov et al., 2014; Lou et al., 2018), the PCDR-based assay demonstrated higher efficiency and at least a ten-fold increased sensitivity than that of the PCR-based assay (Fig.1 and Table 1).

LAMP is a fast and specific isothermal method for DNA amplification. Unfortunately, the use of LAMP for SARS-CoV-2 detection provides robust detection of the virus only if a few hundred (or more) copies of viral RNA are present in the reaction assay (Gray et al., 2020; Zhang et al., 2020). Lower amounts of viral RNA generate a large proportion of false negative results (Ben-Assa et al., 2020); thus, the improvement of assay sensitivity is very important. In an attempt to improve sensitivity, we combined LAMP and PCR and added several PCR cycles before the isothermal amplification. Our hypothesis was that in the case of a low amount of cDNA, the additional PCR cycles would provide amplification of the starting material to the level of DNA copies, which would be sufficient to ensure the start of LAMP. For example, 6 PCR cycles can amplify 5 copies of a target DNA up to 320 copies, which is sufficient for the fast and specific initiation of LAMP.

The results of the RT-qLAMP and RT-q(PCR-LAMP) assays (Figure 2 and Table 2) demonstrated that RT-q(PCR-LAMP) was approximately 100-fold more sensitive than conventional RT-qLAMP. The use of RT-q(PCR-LAMP) allowed us to detect as few as 5 copies of SARS-CoV-2 RNA per reaction and required only 35 min for the amplification step, including 6 cycles of PCR.

In conclusion, the hybrid methods of DNA amplification combined with PCR and isothermal amplification, such as PCDR and PCR-LAMP, allowed us to markedly improve the results of SARS-CoV-2 detection in comparison with conventional PCR and LAMP assays. The hybrid methods reduced the testing time and/or increased the sensitivity against standard amplification methods. The described RT-qPCDR and RT-q(PCR-LAMP) methods are fast and very sensitive and can be used for SARS-CoV-2 RNA detection.

## Data Availability

Data available within the article or its supplementary materials

## Author Contributions

VMK and KBI were responsible for obtaining funding for the research. DAV, KAB, VMK, and KBI contributed to conception and design of the study and data curation. DAV, KAB, and KBI were involved in the investigation and methodology development. KAB, VMK, and KBI carried out the formal analysis. EVS, VMK, and KBI validated the results. EVS and KBI supervised the research. KAB, EVS, and KBI wrote the first draft of the manuscript. EVS and KBI contributed to manuscript review and revision. KBI was responsible for obtaining necessary resources and preparing visuals.

## Acknowledgments

We thank Syntol JSC (Moscow, Russia) for support of this project. We would also like to thank Editage (www.editage.com) for English language editing.

## Data Availability Statement

The original contributions presented in the study are included in the article/supplementary material, further inquiries can be directed to the corresponding author/s.

